# Generating complex explanations for artificial intelligence models: an application to clinical data on severe mental illness

**DOI:** 10.1101/2024.01.09.24300944

**Authors:** Soumya Banerjee

## Abstract

We present an explainable artificial intelligence methodology for predicting mortality in patients. We combine clinical data from an electronic patient healthcare record system with factors relevant for severe mental illness and then apply machine learning.

The machine learning model is used to predict mortality in patients with severe mental illness.

Our methodology uses class-contrastive reasoning. We show how machine learning scientists can use class-contrastive reasoning to generate complex explanations that explain machine model predictions and the data.

An example of a complex class-contrastive explanation is the following: “ The patient is predicted to have a low probability of death because the patient has self-harmed before, and was at some point on medications such as first-generation and second-generation antipsychotics. There are 11 other patients with these characteristics. If the patient did not have these characteristics, the prediction would be different. ”

This can be used to generate new hypotheses which can be tested in follow-up studies.

Our technique can be employed to create intricate explanations from healthcare data and possibly other areas where explainability is important. We hope this will be a step towards explainable AI in personalized medicine.

## Introduction

Artificial intelligence and machine learning have become pervasive in healthcare. Unfortunately many machine learning methods cannot explain why they made a particular prediction. Explainability is very important in critical domains like healthcare.

In this work we show how machine learning scientists can produce complex explanations from data and models. The complex explanations are generated based on class-contrastive reasoning [1] [2]. An example of a class-contrastive explanation is: “This patient is predicted to be in a severely ill category because age is greater than 90 and the patient has a serious disease. If the age was less than 60, then the prediction would have been different”.

In previous work [2], we applied class-contrastive analysis to explain machine learning models. Here we extend the framework and show how machine learning scientists can generate more complex explanations from machine learning models.

We apply these techniques to generate complex explanations from the machine learning model. The machine learning model predicts mortality in patients with severe mental illness (SMI) which is of great public health significance [3]. It is difficult to predict who is at the greatest risk, and therefore who might benefit from interventions and treatments.

Our approach of generating complex explanations can be more broadly applicable in healthcare data and potentially other fields.

## Data and Methods

### Overview of Methods

We give a brief overview of our approach in this section. This is summarised in Fig. 1.

1. We took de-identified data from an electronic patient record system for mental health.
2. We established a collection of features that are not dependent on time, such as age, diagnosis categories, medication categories, and biosocial elements that are essential in severe mental illness (SMI). These features include data on mental health, personal history (e.g., previous suicide attempts and substance abuse), and social predisposing factors (e.g., lack of family support).
3. We used these features to predict death during the time of observation.
4. We then fit machine learning models and classical statistical models.
5. We generated class-contrastive heatmaps to illustrate the explanations of statistical models and machine learning predictions, as well as accompanying statements to facilitate human understanding.
6. Machine learning scientists use these class-contrastive heatmaps to create complex explanations.

**Figure 1.**
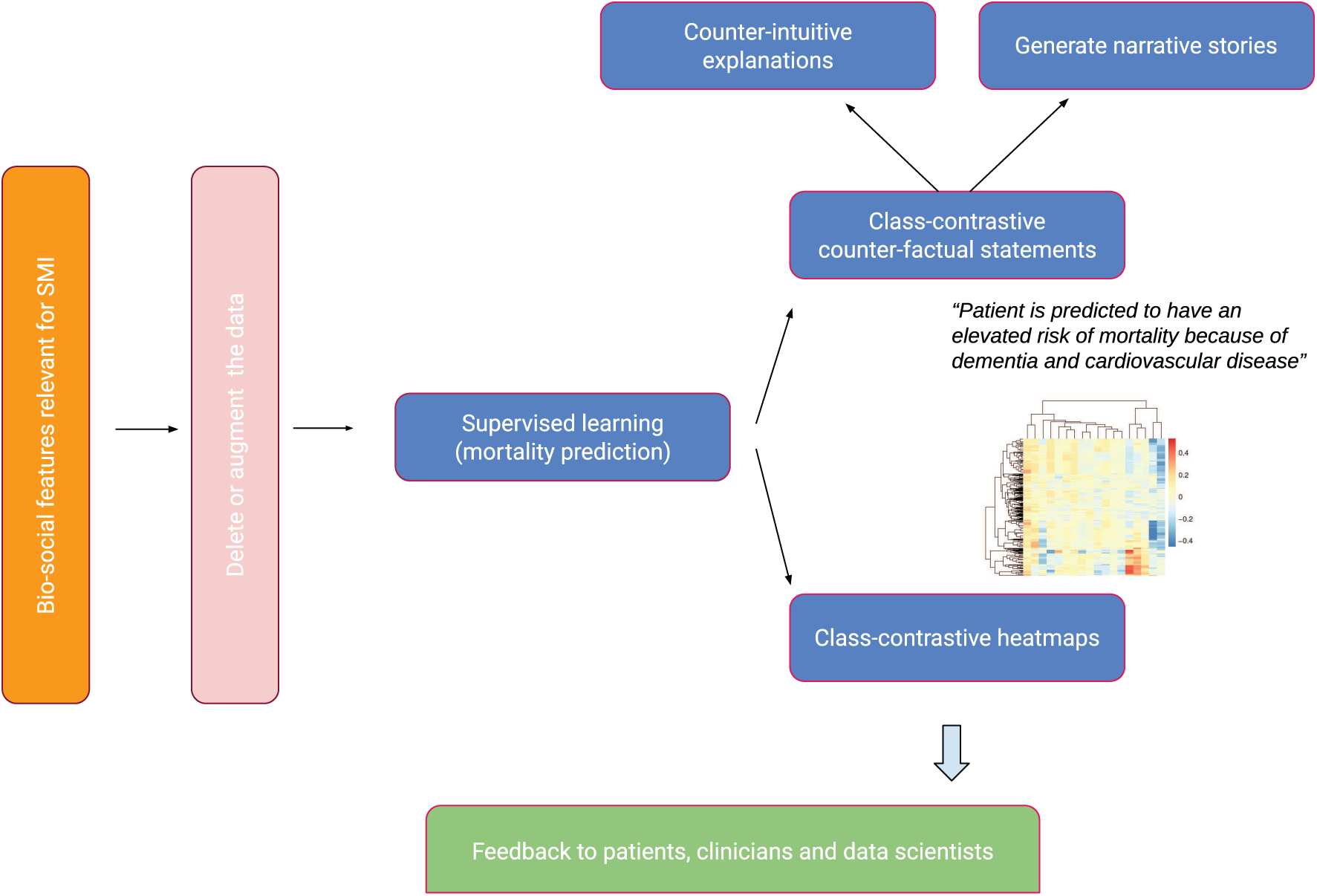
An overview of the approach. Bio-social factors associated with severe mental illnesses (SMIs) come from electronic health records systems and are used to input statistical and machine learning algorithms. These models can be explained with class contradiction reasoning. Machine learning scientists use class-contrastive reasoning to generate complex explanations from data and models. Class-contrastive textual statements and heatmaps help domain experts (such as clinicians, patients, and data scientists) understand models.

We employed artificial neural networks (autoencoders) to predict mortality for patients with schizophrenia. We then constructed a random forest model on top of the autoencoders to predict mortality. The random forest model utilizes the reduced dimensions of the autoencoder as features to predict mortality. We conducted class-contrastive analysis for these machine learning models to make them interpretable.

### Mental health clinical record database

We used data from the Cambridgeshire and Peterborough NHS Foundation Trust (CPFT) Research Database. This comprised of electronic records from CPFT, the single provider of secondary care mental health services for Cambridgeshire. The records were de-identified using the CRATE software [4] under NHS Research Ethics approval (12/EE/0407, 17/EE/0442).

Data included patient demographics, mental health and physical co-morbidity diagnosis (via coded ICD-10 diagnosis and analysis of free text through natural language processing [NLP] tools [5]).

Dates of death were derived from the National Health Service (NHS). There were a total of 1706 patients diagnosed with schizophrenia defined by coded ICD-10 diagnosis. We note that there is under-coding of schizophrenia diagnoses. We included all patients referred to secondary mental healthcare in CPFT from 2013 onwards with a coded diagnosis of schizophrenia.

We extracted medicine information for each patient by using natural language processing (using the GATE software [5] [6]) on clinical freetext data.

### Data input to statistical algorithms

The features that were used as input to our statistical and machine learning algorithms were age, gender, social predisposing factors, high-level diagnosis categories and high-level medication categories. These included a range of bio-social factors that are important in SMI. All these features are used to predict mortality. The full list of features was as follows:

1. High-level medication categories were created based on domain-specific knowledge from a clinician. These medication categories are: second-generation antipsychotics (SGA: clozapine, olanzapine, risperidone, quetiapine, aripiprazole, asenapine, amisulpride, iloperidone, lurasidone, paliperidone, sertindole, sulpiride, ziprasidone, zotepine), first-generation antipsychotics (FGA: haloperidol, benperidol, chlorpromazine, flupentixol, fluphenazine, levomepromazine, pericyazine, perphenazine, pimozide, pipotiazine, prochlorperazine, promazine, trifluoperazine, zuclopenthixol), antidepressants (agomelatine, amitriptyline, bupropion, clomipramine, dosulepin, doxepin, duloxetine, imipramine, isocarboxazid, lofepramine, maprotiline, mianserin, mirtazapine, moclobemide, nefazodone, nortriptyline, phenelzine, reboxetine, tranylcypromine, trazodone, trimipramine, tryptophan, sertraline, citalopram, escitalopram, fluoxetine, fluvoxamine, paroxetine, vortioxetine and venlafaxine), diuretics (furosemide), thyroid medication (drug mention of levothyroxine), antimanic drugs (lithium) and medications for dementia (memantine and donepezil).
2. Relevant co-morbidities we included were diabetes (inferred from ICD-10 codes E10, E11, E12, E13 and E14 and any mentions of drugs metformin and insulin), cardiovascular diseases (inferred from ICD-10 diagnoses codes I10, I11, I26, I82, G45 and drug mentions of atorvastatin, simvastatin and aspirin), respiratory illnesses (J44 and J45) and anti-hypertensives (mentions of drugs bisoprolol and amlodipine).
3. We included all patients with a coded diagnosis of schizophrenia (F20). For these patients with schizophrenia, we also included any additional coded diagnosis from the following broad diagnostic categories: dementia in Alzheimer’s disease (ICD-10 code starting F00), delirium (F05), mild cognitive disorder (F06.7), depressive disorders (F32, F33) and personality disorders (F60).
4. We also included relevant social factors: lack of family support (ICD-10 chapter code Z63) and personal risk factors (Z91: a code encompassing allergies other than to drugs and biological substances, medication noncompliance, a history of psychological trauma, and unspecified personal risk factors); alcohol and substance abuse (this was inferred from ICD-10 coded diagnoses of Z86, F10, F12, F17, F19 and references to thiamine which is prescribed for alcohol abuse). Other features included are self-harm (ICD-10 codes T39, T50, X60, X61, X62, X63, X64, X78 and Z91.5), non-compliance and personal risk factors (Z91.1), referral to a crisis team at CPFT (recorded in the electronic healthcare record system) and any prior suicide attempt (in the last 6 months or any time in the past) coded in structured risk assessments.

These broad categories constituted our domain expert (clinician) based knowledge representation. We used these features to predict whether a patient died during the time period observed (from first referral to CPFT to the present day). We note that we do not attempt to predict the risk of dying in a specific time period, for example, in the next 1 year or in the next 90 days.

These data structures capture different information about the patient related to mental health, physical health, social factors and predisposing factors.

### Data pre-processing

Diagnosis codes were based on the International Classification of Diseases (ICD-10) coding system [7]. Age of patients was normalised (feature scaled). All categorical variables like diagnosis and medications (described above) were converted using a one-hot encoding scheme. This is explained in detail in the Supplementary Section.

### Machine learning and statistical techniques

We used age (feature scaled) as a continuous predictor. There are a large number of categorical features (medications, co-morbidities and other social and personal predisposing factors) which are encoded using a one-hot representation.

We performed logistic regression using generalized linear models [8] [9]

We also used artificial neural networks called autoencoders to integrate data from different sources giving a holistic picture of mental health, physical health and social factors contributing to mortality in SMI [2].

Artificial neural networks are composed of computational nodes (artificial neurons) that are connected to form a network. Each node carries out a basic computation (like logistic regression). The nodes are arranged in layers. The input layer takes in the input features, transforms them and passes them to one or more intermediate layers known as hidden layers. The output layer is used to generate a prediction (mortality).

An autoencoder is a type of artificial neural network that reduces the number of neurons in the hidden layer compared to the input layer, thus performing dimensionality reduction. In our framework, the output of the hidden layer of the autoencoder is used as input to a random forest model (Fig. 2).

**Figure 2.**
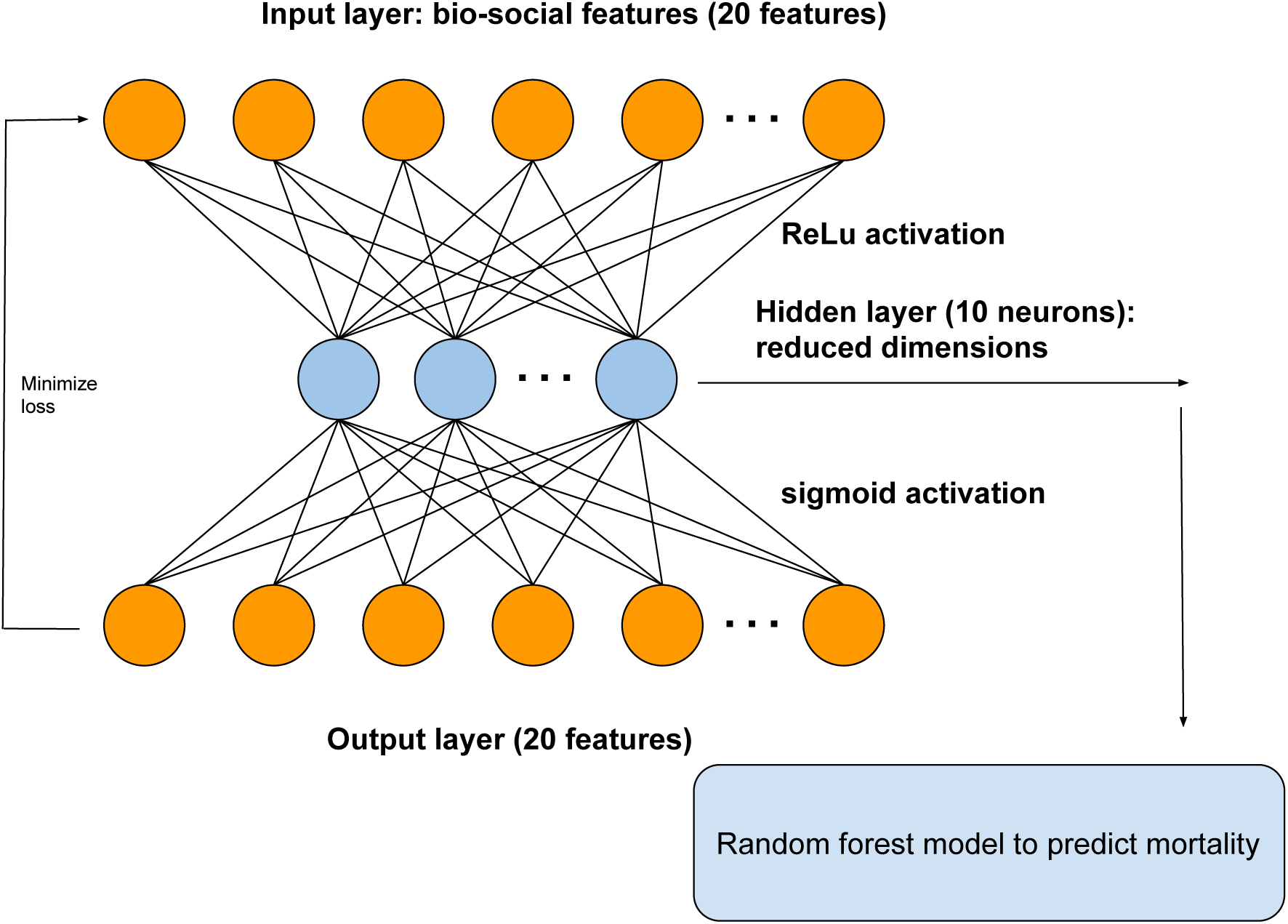
A diagram showing the architecture of autoencoder. The autoencoder takes as input the bio-social features relevant for severe mental illness (SMI). The output layer is used to reconstruct the input. The hidden layer of the autoencoder is used for dimensionality reduction and as input to a random forest model to predict mortality.

Random forests are machine learning models that build collections of decision trees [10]. Each decision tree is a series of choices based on the features. These decision trees are combined to build a collection (forest) that together has better predictive power than a single decision tree.

We performed a 50%-25%-25% training-test-validation split of the data. We performed 10-fold cross-validation with regularization to penalize for model complexity. Additional details are available in Section Machine learning methods and summarised in Fig. 2.

To predict mortality, we used the following models:

1. logistic regression model with all original input features.
2. all the original input features used as input for an autoencoder. The reduced dimensions from an autoencoder were then used as input to a random forest model (Fig. 2).

Class contrastive reasoning

We use class-contrastive reasoning and class-contrastive heatmaps [2] to make our models interpretable. The process involves training the model on the training set. For each patient in the test set, we alter each categorical feature (or a set of features) from 0 to 1 or yes to no. We then measure the change in the predicted mortality probability of the model (on the test set). Predictions are made using the trained machine learning model on the test set.

We repeat this procedure independently for each feature and each patient. We do not retrain the model when we mutate the features. The change in probability is then visualized as a heatmap (class-contrastive heatmap).

In a class-contrastive heatmap, rows represent patients and columns represent the combination of features that are altered at the same time.

We note that age is a factor (predictor) in all the models we use; however, the class-contrastive heatmaps do not take this into account. This is because the class-contrastive analysis only works with categorical features, and so it is not possible to modify age. Therefore, the class-contrastive heatmaps demonstrate the effect of changing features on the model’s predicted probability of mortality, in addition to the contribution of age.

### Machine learning methods

We used artificial neural networks to predict mortality in patients with severe mental illness, specifically schizophrenia. We used an autoencoder to integrate data from different sources giving a holistic picture of mental health, physical health and social factors contributing to mortality in SMI [2].

The input features were age (normalised), gender, diagnosis categories, lifestyle risk factors, social factors and medication categories. Categorical features (like medication categories) were encoded using a one-hot representation. This involves taking a vector that is as long as the number of unique values of the feature. Each position on this vector corresponds to a unique value taken on by the categorical feature. Whenever a categorical feature takes on a particular value (say True), we place a 1 (hot) corresponding to that position on the vector and 0 elsewhere.

We show the architecture of the autoencoder in Fig. 2. The autoencoder has one hidden layer of 10 neurons. The autoencoder takes as input the bio-social features. The output layer is then used to reconstruct the input. The hidden layer of the autoencoder is used for dimensionality reduction. We also used the hidden layer as input to a random forest model.

We performed a 50%-25%-25% training-test-validation split of the data. The autoencoder used a Kullback-Leibler loss function (discussed below). We used the *keras* package [11] with the *Tensorflow* backend [12].

An artificial feed-forward neural network optimizes a loss function of the form:

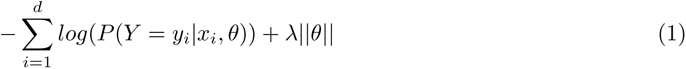

This is the negative log-likelihood. There are *d* data points. The *i* th data point has a label denoted by *y_i_* and input feature vector represented by *x_i_*. The weights of the artificial neural network are represented by a vector *θ*. *λ* is a regularization parameter to prevent overfitting and reduce model complexity. *λ* is determined by 10-fold cross-validation. Shown here is the *L*_1_ norm of the parameter vector.

An activation function is used to project the input matrix (*X*) into another feature space using weights (*W* ). These weights are determined using backpropagation.

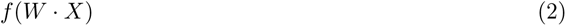

We used a ReLU (Rectified Linear Unit) activation function for the hidden layers. The form of the ReLU function is shown below:

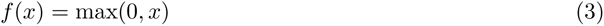

We added an *L*_1_ penalty term for the weights to perform regularization and performed 10-fold cross-validation to select the regularization parameter (*λ*).

The autoencoder used a Kullback-Leibler loss function, which is a measure of discrepancy between the input layer and reconstructed hidden layer.

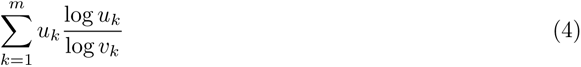

where there are *m* features in the input layer. *u* represents the input layer and *v* represents the hidden layer. The layers are calculated by applying the appropriate activation functions.

We also experimented with a categorical-cross entropy loss function for the autoencoder (shown below) [13]. However our results were not changed significantly.

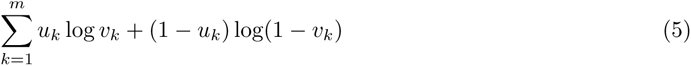

The final cost function is given below:

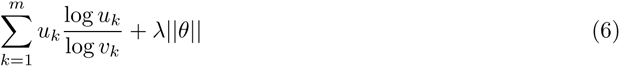

where the vector *θ* represents all the weights of the artificial neural network. There are *m* features in the input layer. *u* represents the input layer and *v* represents the hidden layer. *λ* is a regularization parameter which prevents overfitting.

We initialized weights of the artificial neural network according to the Xavier initialization scheme [14] and used the *Adadelta* method of optimization [15]. An artificial neural network is trained on the training data for a number of epochs. Our neural networks were trained for 1000 epochs, which was assessed as being sufficient enough to reach convergence.

### Software

All software was written in the R [16] and Python programming languages. The deep learning algorithm was implemented using the *keras* package [11] with the *Tensorflow* backend [12]. Logistic regression was performed using the *glm* function in R [17] [9]. Heatmaps were visualized using the *pheatmap* package [18]. All our software is freely available here:

https://github.com/neelsoumya/machine_learning_advanced_complex_stories

Code to perform similar analysis on a publicly available dataset is available here:

https://github.com/neelsoumya/complex_stories_explanations

## Results

### Class-contrastive reasoning and modifying the training data to generate explanations

A machine learning algorithm can be explained by both how the model itself explains predictions and how it uses the idiosyncrasies in the data to derive explanatory power. We seek to answer the question: from which training data examples does the ability to make a particular prediction come from? We remove specific training data examples to answer this question.

In the first instance, we use class-contrastive reasoning on a logistic regression model. The logistic regression model is trained on the training data. We then change one feature at a time on the test data and record the change in the model prediction (on the test set). In this scenario, the model is not retrained. The change in model predictions is visualized as a class-contrastive heatmap.

A class-contrastive heatmap is shown in Fig. 3. This heatmap suggests some intuitive and counter- intuitive statements.

**Figure 3.**
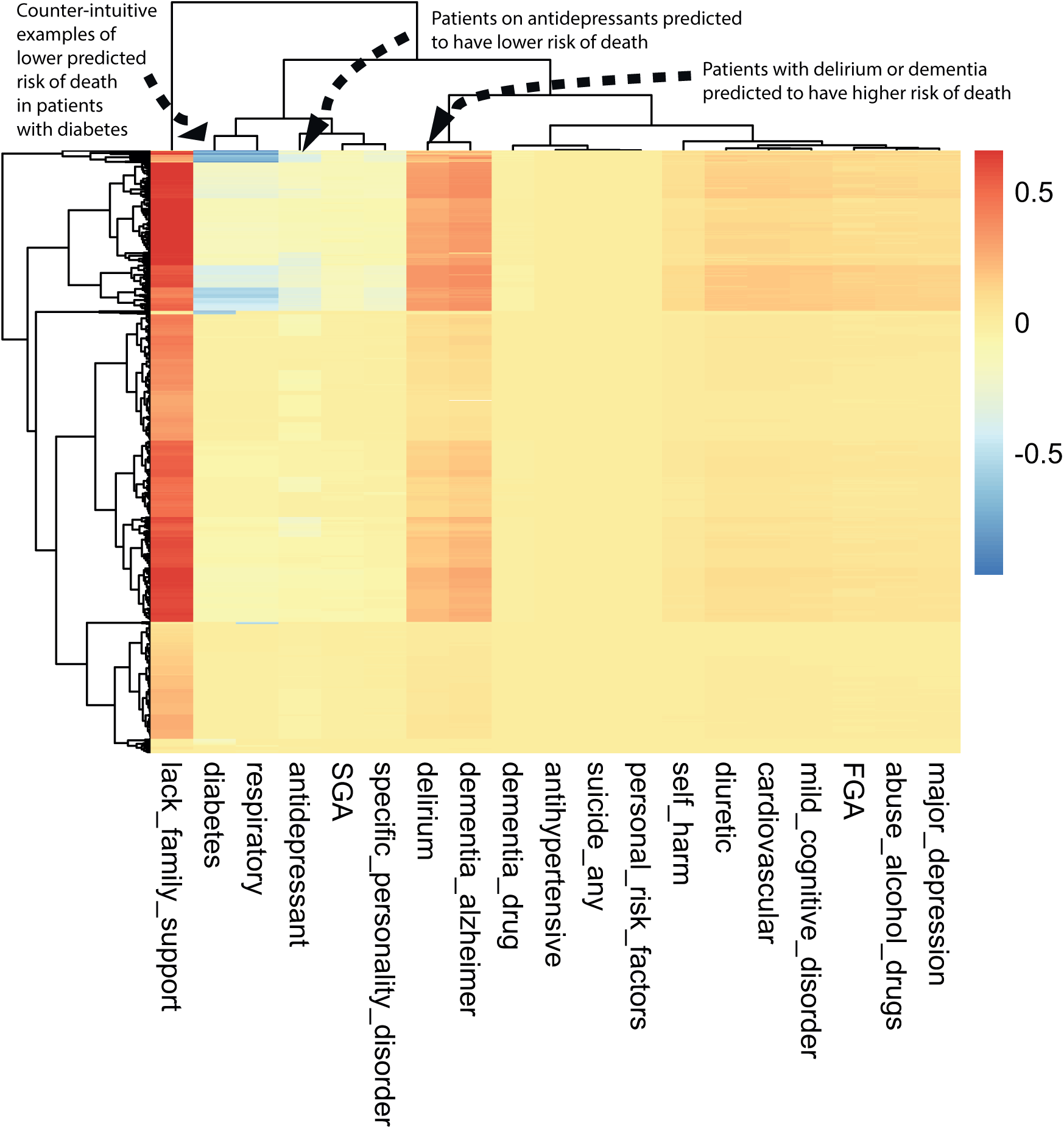
A visualization of the amount of change predicted in the probability of death by setting a particular feature to 1 versus 0 (using a logistic regression model) on the test set. The rows represent patients and the columns represent features. The predictions are made using a logistic regression model on the test set. This class-contrastive heatmap also shows a hierarchical clustering dendrogram performed using an Euclidean distance metric and complete linkage. The arrows show groups of patients with low predicted risk of mortality (shown in blue on the heatmap) using the logistic regression model. The arrow at the top left shows another group of patients taking antidepressants. These patients are predicted (using the logistic regression model) to have a lower risk of mortality. The arrow at the top left corner shows a group of patients having diabetes and still have low predicted risk of death: this is counter-intuitive.

We observe that a diagnosis of delirium or dementia seems to predispose a group of patients towards a higher probability of predicted mortality (Fig. 3). Additionally the class-contrastive heatmap suggests that a group of patients on antidepressants were less likely to die during the period observed (Fig. 3).

One counter-intuitive class-contrastive statement is: “Use of or prescription of second-generation antipsychotics (SGA) and any prior suicide attempt is correlated with a decline in mortality”. This is a very counter-intuitive statement. To get at why such a counter-intuitive statement was produced, we seek to understand which parts of the training data influenced the model to come up with this statement. The data and the model are entangled. In order to disentangle this, we do the following:

1. we use case-based reasoning to determine which patients are similar and have these characteristics.
2. we then remove these data from the training set.
3. we retrain the statistical model on this modified data.

We hypothesized that the statistical model is learning a pattern from the data since there is a subset of patients who are alive, have used SGA and have attempted suicide. We removed all instances of co-occurrence of patients being alive, having attempted suicide, and also used or being prescribed SGA (prior suicide attempt = 1 and SGA = 1 and death flag = 0). There were 539 patients with these characteristics which we removed. This is similar to case-based reasoning since we identified 539 cases that are similar to each other. This left us with 1167 patients with schizophrenia.

We then fit a logistic regression model on this modified data. The log-odds ratio from the model is shown in Fig. 4. On this modified data, prior suicide attempt has a log-odds ratio greater than 1 (Fig. 4). Hence the apparent association of prior suicide attempts with lower mortality was removed after modifying the training data.

**Figure 4.**
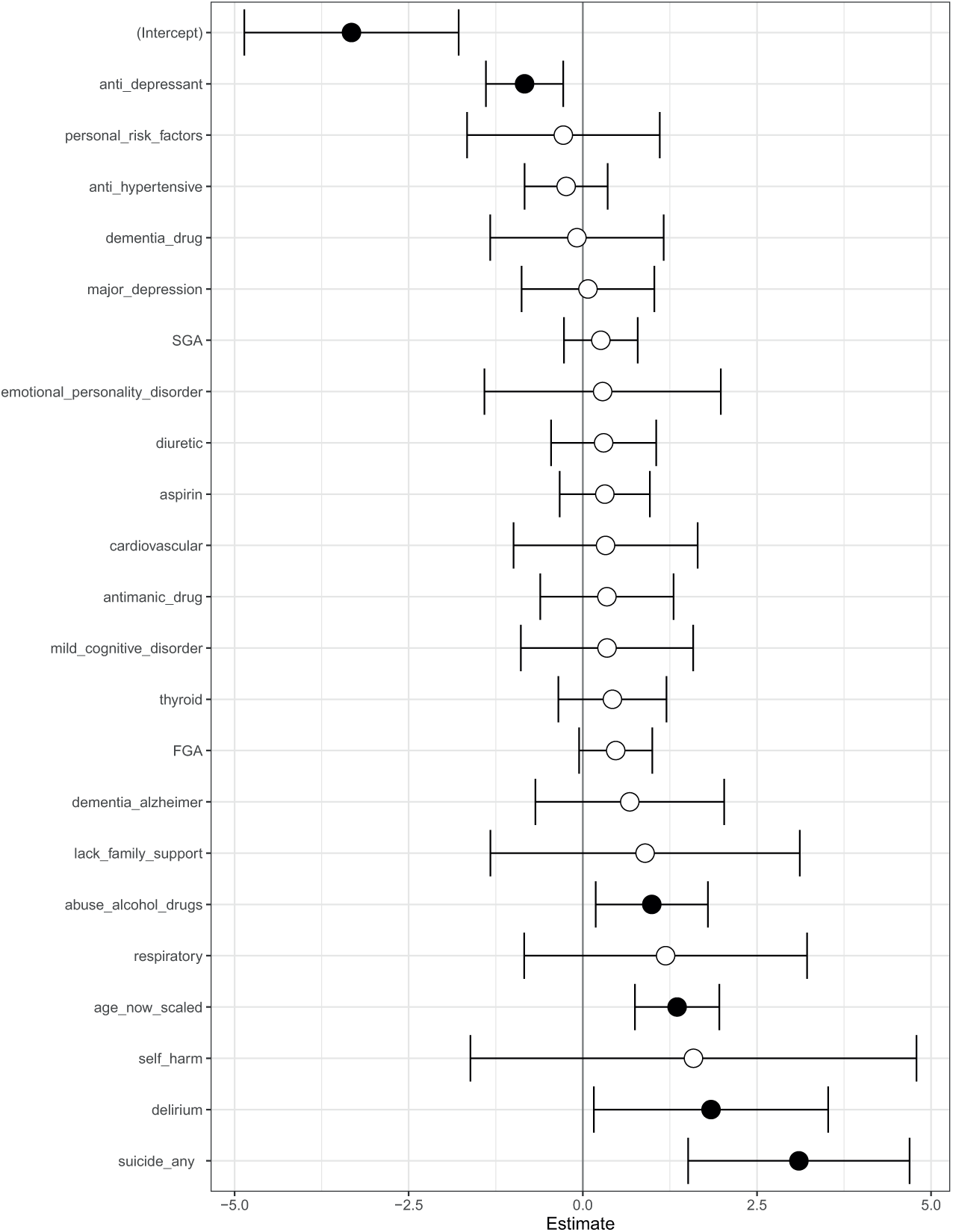
A visualization of the log-odds ratios from a logistic regression model trained on the modified data where all co-occurrence of prior suicide, SGA use and death have been removed. Shown are confidence intervals and statistical significance (filled dark circles: p-value *<* 0.05, open circles: not significant).

This suggests that there may be a subset of patients with schizophrenia where prior suicide attempt is associated with reduced mortality. It maybe that after attempted suicide these patients have received very good care from mental health service providers. This is subject to verification in future follow-up studies in an independent cohort with more patients. Our objective here is only to raise hypotheses in an observational study that can be verified in follow-up studies.

We suggest that this data-centric approach can be used to explain machine learning models. This can complement the purely model-centric approach that has dominated techniques for explainable AI.

The counter-intuitive observations made on the test set can also be the result of imbalances in the training set. For example, a particular binary feature can be 1 for 1000 patients and 0 for 10 patients.

Finally, we note that age is a factor (predictor) in all the models we use; however, the class-contrastive heatmaps do not take this into account. This is because the class-contrastive analysis only works with categorical features, and so it is not possible to modify age. Therefore, the class-contrastive heatmaps demonstrate the effect of changing features on the model’s predicted probability of mortality, in addition to the contribution of age.

### Class contrastive reasoning applied to machine learning models

We can also use this class-contrastive reasoning technique to explain machine learning models [2]. The machine learning model is trained on the training data and then used to predict mortality. We then take into account all pairs of features in the test set: for each pair of features in the test set, we then simultaneously changed the values from 0 to 1. We record the difference in the probability of mortaility (predicted using the model on the test set). This process is then repeated for each pair of features and for each patient on the test set.

We then visualize the change in the model predicted probability of mortality (produced by the trained machine learning model in the now modified test set). We visualize this using a heatmap, which we call a class-contrastive heatmap (Fig. 5).

**Figure 5.**
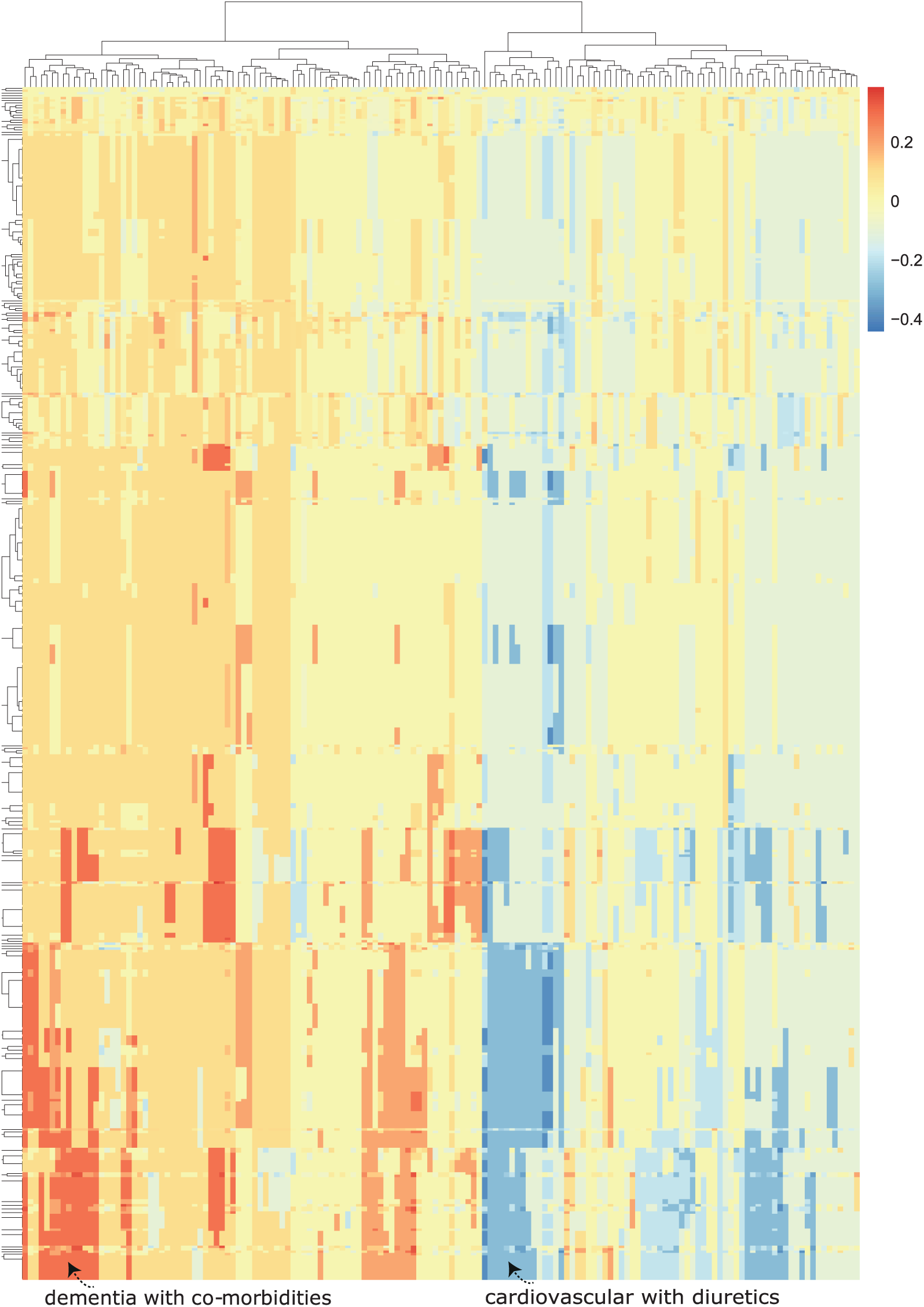
A class contrastive heatmap for the deep learning model showing the effect of interactions between features. This is a visualization of the amount of change predicted in the probability of death (from the deep learning model) by setting a particular combination of two features to 1 simultaneously (versus 0) on the test set. The rows represent patients and the columns represent groupings of features (all combinations of two features). The predictions are made on the test set using a deep learning model. Delirium and dementia in Alzheimer’s disease seem to increase the risk of death for some patients: this is shown in red in the lower left-hand corner of the heatmap. Diuretics appear to be associated with lower probability of predicted mortality in some patients with cardiovascular disease: see the blue region at the lower right-hand corner of the heatmap). The heatmap also shows a hierarchical clustering dendrogram: this is created using an Euclidean distance metric and complete linkage.

This heatmap shows the importance of combinations of features. For example, diuretics seem to be associated with a lower probability of mortality (as predicted by the machine learning model) in a group of patients with cardiovascular disease. This is shown in the blue region of the heat map at the bottom right-hand corner (Fig. 5). This is the region where the predicted probability of death is reduced most. The combination of delirium and dementia in Alzheimer’s disease may also predispose some patients towards a higher probability of predicted mortality (as predicted by the machine learning model on the test set). This is shown in the lower left region of the heatmap in red (Fig. 5).

### Generating complex explanations

We then applied class-contrastive reasoning to the deep learning model, where now we simultaneously changed three features on the test set from being set to all 1 to being set to all 0. We consider all combinations of three features on the test set. For a particular combination of three features, we set all of them simultaneously to 1. We then record the model predicted probability of mortality (for all patients on the test set). We then set those three features simultaneously to 0 and record the model predicted probability of mortality (for all patients on the test set). The difference in these two probabilities is also stored. This process is then repeated for all combinations of three features.

The rows of a class-contrastive heatmap represents patients and the columns represent those above-mentioned difference in probabilities. A class-contrastive heatmap for this is shown in Fig. 6.

**Figure 6.**
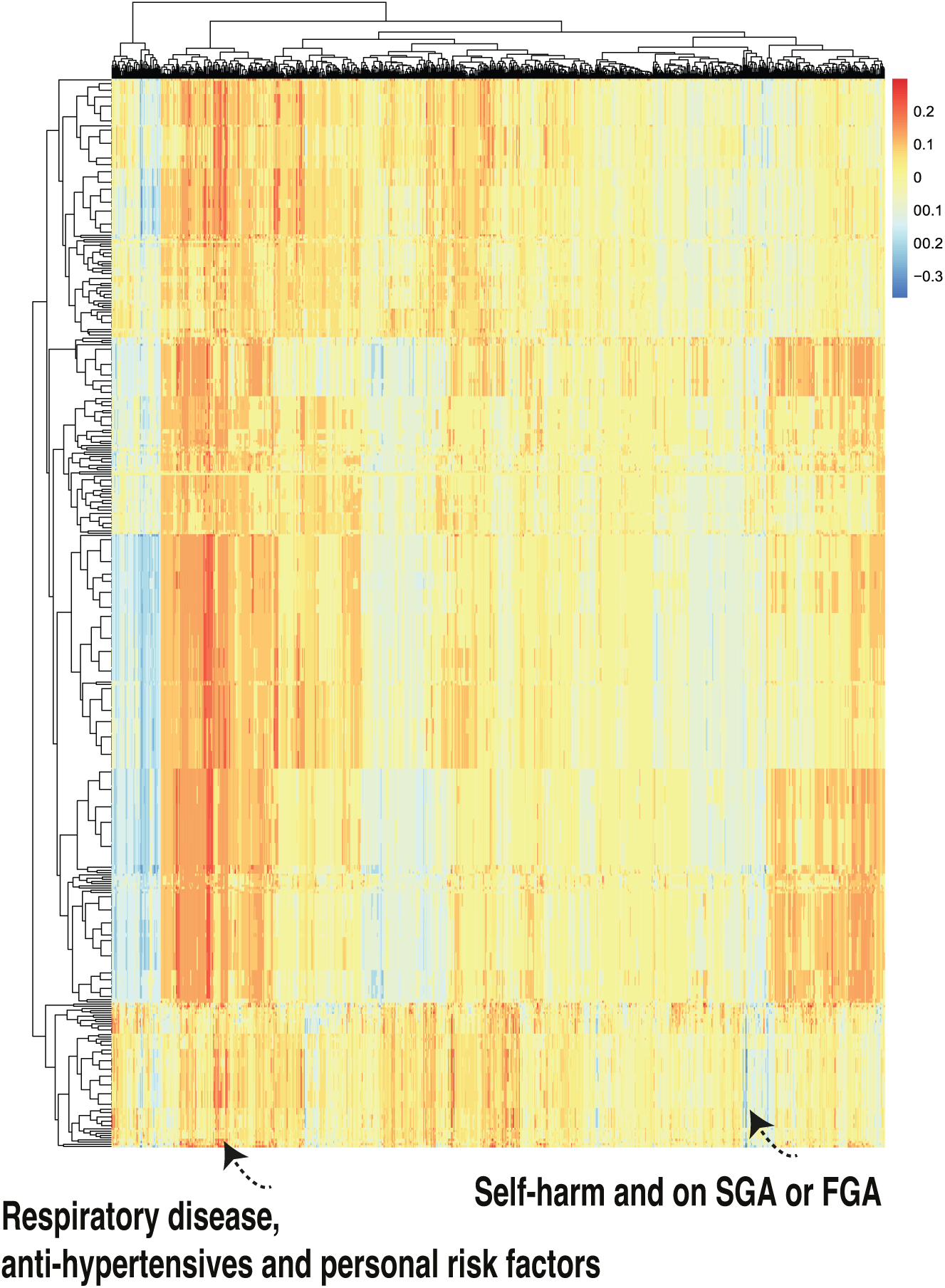
A class contrastive heatmap for the deep learning model where 3 features are changed from 1 to 0 simultaneously on the test set. The heatmap shows the amount of change predicted in the probability of death by setting 3 combinations of features simultaneously to 1 versus 0. The rows represent patients and the columns represent all combinations of 3 features. The predictions are made with a deep learning model on the test set. The figure shows two groups of patients. The group in the left-hand corner is composed of patients with respiratory disease, who are taking anti-hypertensives and have personal risk factors: they are predicted by the machine learning model to be at higher risk of mortality. The group in the right-hand corner of the heatmap is composed of patients who have self-harmed and were at some point on second-generation antipsychotic (SGA) or first-generation antipsychotic (FGA) medications. Counter-intuitively this group of patients is predicted (by the machine learning model) to have lower risk of mortality.

There is one patient (Fig. 6, right hand corner) who is predicted to have a reduced probability of mortality. The class-contrastive techniques suggests that if the patient self harms and was at some point on SGA or FGA, this would lead to a lower risk of predicted mortality (as predicted by the machine learning model on the test set).

Using case based reasoning we found 11 cases that are similar to each other (have self-harmed and was at some point on SGA and FGA). There are 11 such patients of whom 1 has died. We hypothesize that the deep learning algorithm may be inferring that the simultaneous co-occurrence of these features is associated with protection (against mortality).

The class-contrastive explanation here is: “The patient is predicted to have a low probability of death because the patient has self-harmed before, and was at some point on SGA or FGA. There are 11 other patients with these characteristics. If the patient did not have these characteristics, the prediction would be different.”

Hence we can build ever more complex explanations and narratives using class-contrastive reasoning. These narratives can be used to build hypotheses which can be tested in future studies.

We complemented this class-contrastive analysis by fitting a simple logistic regression model with all main effects and an interaction effect between self-harm and SGA fit to the same data (Fig. 7). We observe that self-harm interacts with FGA (p-value = 0.03). It is possible that patients who have self-harmed are put on FGA and the medication has a protective effect. We note that our objective is only to raise novel hypotheses which will ultimately need to be tested in future studies.

**Figure 7.**
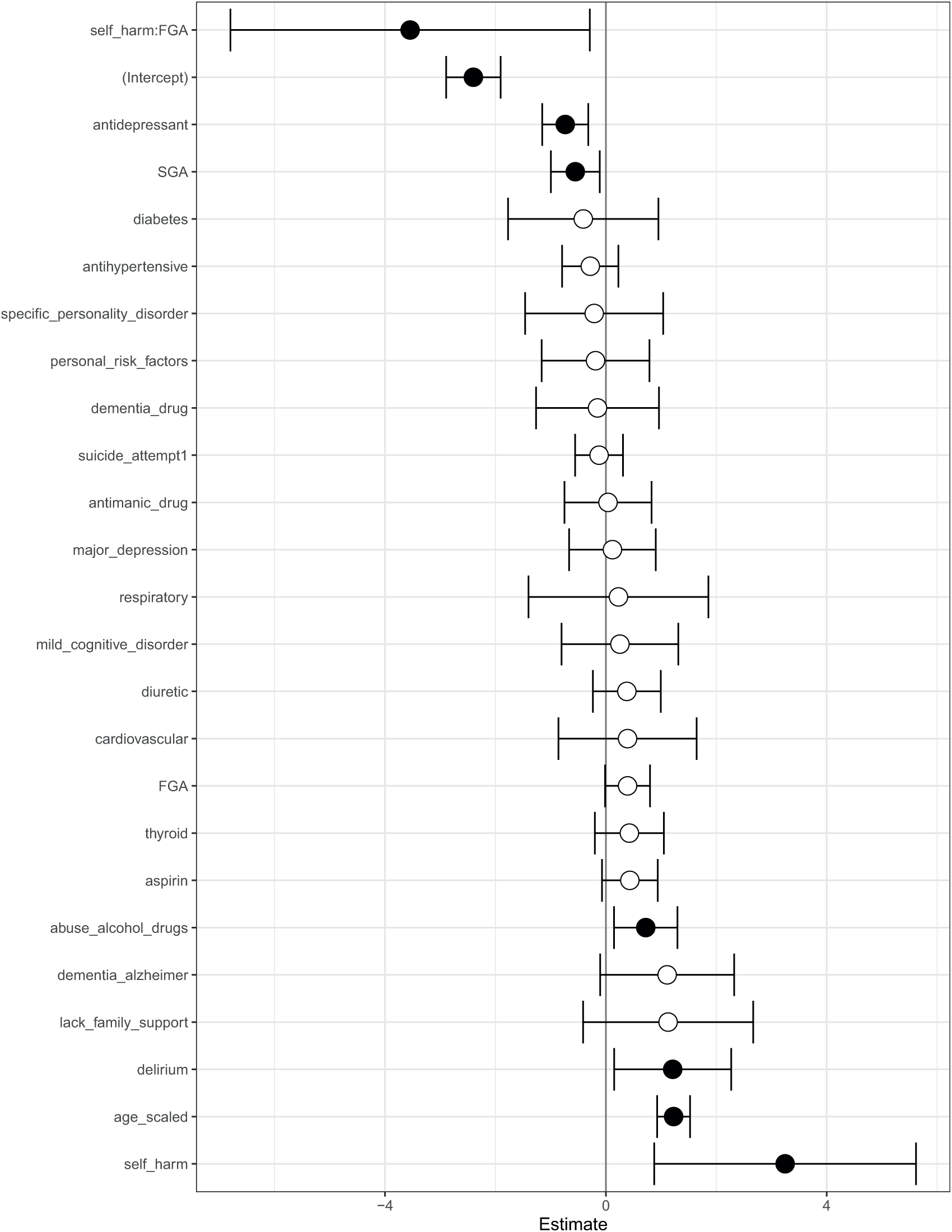
Odds ratios from a logistic regression model to predict mortality. We show the main effects and an interaction effect between self harm and use of first-generation antipsychotics (FGA). Also shown are the odds ratios corresponding to use of FGA and second-generation antipsychotics (SGA). Statistical significance is denoted by filled dark circles (p-value *<* 0.05) and open circles (not significant).

There is an imbalance between the number of patients who have self-harmed, are on SGA, and also on FGA (11 patients) and the number of patients who have not self-harmed, are not on SGA and also not on FGA (152 patients). There are very few patients in one of these groups (11 patients). This will have implications for inferring parameters in the machine learning and logistic regression model. Hence these counter-intuitive narratives may be generated because the machine learning algorithm was not able to understand the real importance of the combinations of the features since they were very rare in the data. We explore this in more detail in Section Counter-intuitive narratives.

We note that our objective is to raise hypotheses and not posit novel mechanisms or generate novel clinical insights. Our approach can be used to generate new hypotheses which can be tested in follow-up studies with more data or interventional studies (randomized control trials).

### Counter-intuitive narratives

Our approach is not without limitations. Machine learning scientists can used class-contrastive reasoning to generate increasingly complex explanations. However, in some instances this can lead to explanations that are counter-intuitive.

As progressively more complex stories are generated, there is less data to perform inference on these complex and unique patterns. We hypothesize that this leads to counter-intuitive stories. These counter- intuitive statements (or counter-factual narratives) can also be adversarial examples and be generated due to spurious learning [19]: the machine learning model picking up spurious statistical patterns in the data. These may point to additional data collection efforts or motivate new hypotheses.

We outline such an example here. One complex explanation is: “ There are 3 patients with respiratory disease, on anti-hypertensives and having personal risk factors: they are predicted to be at a higher risk of mortality”. This is shown in Fig. 6 (left hand corner). However there were no patients in either the training set or test set with these features i.e. have respiratory disease, are on anti-hypertensives and have personal risk factors. Hence the machine learning algorithm may have picked up a spurious pattern.

We hypothesize that these counter-intuitive stories can lead to contradictions but they might also suggest follow-up studies and further investigations. Hence this may be used to generate hypotheses which can be tested using additional data.

## Discussion

### Overview

Clinical data collected routinely in hospitals can help generate insights. We used a machine learning model to predict the mortality of patients with SMI using clinical data collected regularly. We present an approach where machine learning scientists can generate complex explanations of the machine learning model.

Using clinical information on physical, mental, personal and social factors, we create profiles of patients. The machine learning model predicts patient mortality. The predictions are then explained by machine learning scientists by using class-contrastive reasoning. We extend previous work [2] to account for more complex explanations of the data and the model.

We generate complex explanations to explain specific features of the data and the model. We use these as a mechanism to generate hypotheses, which can be investigated in follow-up studies with more patients.

### Implications of findings

The methodology described here can be used by machine learning scientists to visually inspect the class-contrastive heatmaps and create complex explanations. These can be used to generate new hypotheses and design new follow-up studies.

Our class-contrastive framework highlights some counter-intuitive results. One counter-intuitive statement is: “Features which if set to 0 provide most positive change in probability of predicted mortality are: any prior suicide attempt and use of SGA”

We hypothesized that the machine learning model is learning a pattern from the data since there is a subset of patients who are alive, and have been prescribed SGA and have attempted suicide. We removed all instances of co-occurrence of suicide attempt and prescription of SGA and the patient was alive i.e. suicide = 1 and SGA = 1 and death flag = 0. There were 539 patients with these characteristics, which we removed. This is similar to case-based reasoning since we identified 539 cases that are similar to each other. This left us with 1167 patients.

We then fit a logistic regression model on this modified data (Fig. 4). Now prior suicide attempt has a log-odds greater than 1. This now takes away the apparent association of prior suicide with reduced mortality.

This suggests there may be a subset of patients where suicide attempt is associated with reduced mortality. It maybe that after attempted suicide these patients have received very good care from mental health service providers. This is subject to verification in future follow-up studies. Our objective is only to raise hypotheses in an observational study that can be verified in follow-up studies.

Diuretics seem to be associated with a lower probability of mortality (as predicted by the machine learning model) in a group of patients with cardiovascular disease. This is shown in the blue region of the heat map at the bottom right-hand corner (Fig. 5). The combination of delirium and dementia in Alzheimer’s disease may also predispose some patients towards a higher probability of predicted mortality (as predicted by the machine learning model on the test set). This is shown in the lower left region of the heatmap in red (Fig. 5).

### Relevant work

Our approach uses elements of case-based and analogy-based reasoning [20]. Analogy based reasoning may be central to how humans generate insights into problems [21].

Many deep learning approaches can also pick up spurious statistical patterns in the data [19]. We note that it is possible that many of the patterns so discovered are spurious. Our findings will need to be validated in future studies with more patient data and may also serve as adversarial examples. Our approach may help uncover weaknesses in learning spurious patterns from data using deep learning models.

There have also been efforts to characterize what kinds of shortcuts are being taken by machine learning algorithms [22] [19]. Our approach may also help uncover shortcut learning in artificial neural networks.

The complex explanations that we suggest are similar to stories. Telling, understanding and recombining stories is a powerful component of human intelligence [23] [24]. Story generating systems are thought to underlie human intelligence [23]. Stories may help humans learn to generalize and specialize from small numbers of examples.

Story generating systems are a form of symbolic AI and are an evolution from earlier efforts to integrate symbols and logic in AI. Symbolic AI is thought to be a really important component of future AI systems that can reason [25] [26].

We present an approach where machine learning scientists can work collaboratively with machines to generate stories from data and models. Our approach is a step towards computationally generating stories as explanations from machine learning models.

## Limitations and future work

We use observational data from a clinic. We do not imply causation. Hence our results should not be used to change clinical practice. Our objective is to raise hypotheses that will need to be tested in further studies with more patients or randomized controlled trials.

In this work, we generated the narratives based on a visual inspection of the class-contrastive heatmaps. This is subjective and may lead to bias. In future work, we will look at computationally building narratives. Computational techniques can also be used to explain disagreements or consensus between different kinds of statistical models. In future work, we will also look at automatically generating explanatory stories from data [1] [27] [2] which can engage humans [24] [23] [28].

Narratives can also be misleading and reflect cognitive biases. Future work will investigate how to protect against misuse of narratives and remove possible bias in stories. There also needs to be a pluralistic evidence base for generating narratives that accounts for multiple sources of evidence, multiple viewpoints or data sources [29].

In high dimensions there can be many adversarial examples and also many counterfactual explanations. Adversarial examples are similar to class-contrastive or counterfactual explanations. We acknowledge that the class-contrastive examples and the stories we generate can indeed be artefacts of the training process, training data and the model: these could be adversarial examples that have no meaning. Hence these explanations may not represent possible worlds or map to realistic scenarios.

Our approach may also uncover noise. This can lead to counter-intuitive narratives (see Section Counter-intuitive narratives). Distinguishing legitimate discoveries from pure noise will require further studies with more patients.

Confounding effects also complicate explainability. For example, there are also side effects of medications such as SGA which may lead to cardiovascular diseases, which may be subsequently managed using diuretics. Hence our approach may be uncovering these relationships between SGA, cardiovascular disease and diuretic use.

We note that we currently also do not control for multiple comparisons that are made implicitly during the process of generating complex explanations.

Finally, we note that our approach of finding the basis of counter-intuitive predictions and shaping the data, can also used by malicious adversaries to inject the precise kinds of training data needed to change the way a machine learning model behaves.

## Concluding remarks

The nature of explanations is multi-faceted and complex [30]. We focus on class-contrastive techniques to generate complex explanations in a machine learning model. The model is used to predict mortality in patients with SMI.

We show how machine learning scientists can use class-contrastive reasoning to generate complex explanations. As an example of a complex explanation, we show the following: ‘The patient is predicted to have a low probability of death because the patient has self-harmed before, and was at some point on SGA or FGA. There are 11 other patients with these characteristics. If the patient did not have these characteristics, the prediction would be different.”

A class-contrastive statement for one randomly selected patient is: “The selected patient is at high-risk of mortality because the patient has dementia in Alzheimer’s disease and has cardiovascular disease. If the patient did not have both of these characteristics, the predicted risk will be much lower”.

We can build ever more complex explanations using class-contrastive reasoning. These complex explanations can be used to build hypotheses which can be tested in future studies. We think of our approach as a hypothesis generating mechanism rather than an aid for decision making for clinicians.

The complex explanations generated by us are similar to stories. Stories have the ability to engage humans [23] [24]. Story generating systems potentially also underlie human intelligence [23]. The work presented here can be used to computationally build narratives that explain data and models [24] [31] [32]. Data scientists can use human-in-the-loop computational techniques to generate explanatory stories and narratives from data and models.

Our work combines classical approaches to AI with deep learning. Approaches like these that combine the strengths of deep learning with traditional AI approaches (like structured rule-based representations, case-based reasoning and commonsense reasoning), are likely to yield promising results in all fields of applied machine learning [33] [34].

Explainability in machine learning models suffers from a number of key limitations [35] [36] [37]. Some of the limitations have to do with the nature of high-dimensional data and the counter-intuitive nature of high-dimensions that the model and data reside in [38] [39]. Others have to do with how counter-intuitive predictions may arise due to idiosyncrasies of the data. These issues are entangled in complex ways. Our method provides a step towards disentangling some of these issues. We note that our method does not completely solve these issues but we hope may provide a fruitful next step towards development of more practical explainable AI techniques.

We note that our aim is not to improve accuracy (with these machine learning techniques) but produce a constellation of interpretable methods: these would range from classical statistical techniques (such as logistic regression) to modern machine learning techniques. More details on accuracy and comparisons of our machine learning technique with other methods are available in [2].

Our framework can also be very useful for clinical decision support systems. These techniques could ultimately be used to build a conversational AI that could explain its predictions to a domain expert. This would need comprehensive evaluation and testing in a clinical setting.

In summary, we present an approach where machine learning scientists interrogate models and build complex explanations from clinical data and models. Our methodology may be broadly applicable to domains where explainability of machine learning models is important.

## Acknowledgements

We thank Rudolf Cardinal and Peter Jones for helpful discussions. We thank Daniel Porter, Jenny Nelder and Jonathan Lewis for their support during this project. This work is dedicated to the memory of Tathagata Sengupta.

## Funding statement

SB acknowledges funding from the Accelerate Programme for Scientific Discovery Research Fellowship. This work was also partially funded by an MRC Mental Health Data Pathfinder grant (MC PC 17213). and was supported by the NIHR Cambridge Biomedical Research Centre (NIHR203312) and the NIHR Applied Research Collaboration East of England. The views expressed are those of the author(s) and not necessarily those of the funders, the NIHR or the Department of Health and Social Care. The funders had no role in study design, data collection and analysis, decision to publish, or preparation of the manuscript.

## Conflicts of interests

SB has no conflicts of interest to disclose.

## Ethics

The CPFT Research Database operates under UK NHS Research Ethics approvals (REC references 12/EE/0407, 17/EE/0442; IRAS project ID 237953). All methods were carried out in accordance with relevant guidelines and regulations. All experimental protocols were approved by a named institutional and/or licensing committee. Informed consent was obtained from all subjects and/or their legal guardian(s)

## Data accessibility

This study reports on human clinical data which cannot be published directly due to reasonable privacy concerns, as per NHS research ethics approvals and NHS information governance rules.

## Data availability

The CPFT Research Database is private. This study reports on human clinical data which cannot be published directly due to reasonable privacy concerns, as per NHS research ethics approvals and NHS information governance rules.

## Software availability

All our software is freely available here:

https://github.com/neelsoumya/machine_learning_advanced_complex_stories

Code to perform similar analysis on a publicly available dataset is available here:

https://github.com/neelsoumya/complex_stories_explanations

## Author contributions

SB carried out the analysis and implementation, participated in the design of the study and wrote the manuscript. All authors gave final approval for publication. SB directed the study.

## Notes

### Competing Interest Statement

The authors have declared no competing interest.

### Author Declarations

The CPFT Research Database approved this and operates under UK NHS Research Ethics approvals (REC references 12/EE/0407, 17/EE/0442; IRAS project ID 237953).

### Summary of Updates

supplementary section revised. The section has now been moved to Methods.

